# Autoimmunity to the Lung Protective Phospholipid-Binding Protein Annexin A2 Predicts Mortality Among Hospitalized COVID-19 Patients

**DOI:** 10.1101/2020.12.28.20248807

**Authors:** Marisol Zuniga, Claudia Gomes, Steven E. Carsons, Michael T. Bender, Paolo Cotzia, Qing Robert Miao, David C. Lee, Ana Rodriguez

## Abstract

**Background:** Annexin A2 is a phospholipid-binding protein involved in fibrinolysis, cell membrane stabilization and repair, and ensuring the integrity of the pulmonary microvasculature. Given the autoantibodies observed in COVID-19 and that Annexin A2 is a known target of antiphospholipid antibodies, we studied autoimmunity directed against Annexin A2 among hospitalized COVID-19 patients.

**Methods:** We used ELISA to identify the levels of IgG autoantibodies recognizing Annexin A2 and A5 among 86 hospitalized cases of COVID-19. Using logistic regression, we analyzed the association between anti-Annexin A2 and A5 antibody levels with mortality after adjusting for age, sex, race and key comorbidities.

**Results:** We found higher average levels of anti-Annexin A2 antibodies among hospitalized COVID-19 patients that died when compared with non-critical hospitalized COVID-19 patients (p-value = 0.006) and critically ill COVID-19 patients (p-value = 0.04). No significant differences in anti-Annexin A5 antibody levels were identified. Regression analysis showed that anti-Annexin A2 antibody levels as measured in relative units strongly predicted mortality with an odds ratio of 9.3 (95% CI: 1.9 to 44.6, p=0.005). In contrast, anti-Annexin A5 antibody levels were not associated with higher mortality (95% CI: 0.5 to 15.2, p=0.22).

**Conclusions:** We determined that anti-Annexin A2 antibodies were elevated among hospitalized COVID-19 patients and these levels predicted mortality. It is known that inhibition of Annexin A2 induces systemic thrombosis, cell death, and non-cardiogenic pulmonary edema. Autoimmunity to Annexin A2 is a potential mechanism that may explain the key clinical findings of severe COVID-19.

## INTRODUCTION

The underlying pathophysiology of the novel coronavirus 2019 (COVID-19) has largely been attributed to a hyper-inflammatory response without a clear indication of the underlying mechanism.(*1*) There has been evidence that autoimmunity may play an important role in the pathogenesis of several conditions associated with COVID-19 such as Guillain-Barre syndrome, autoimmune hemolytic anemia, immune thrombocytopenia purpura, autoimmune encephalitis, and Kawasaki’s disease.(*2*) In addition, recent studies of hospitalized COVID-19 patients have demonstrated that the majority have positive results to some of the most commonly screened antiphospholipid antibodies.(*3*) Some COVID-19 patients have also reported persistent symptoms, many of which could be characterized as being rheumatologic in origin.(*4*)

It remains unknown how some patients who contract the SARS-CoV-2 virus can be asymptomatic whereas others die of severe respiratory failure.(*5*) Pathology reports among COVID-19 deaths demonstrate fibrin and platelet microthrombi in the pulmonary vasculature, diffuse lung damage, and signs of pulmonary edema and fibrin deposition in the alveoli.(*6*) Furthermore, systemic thrombosis in the arterial, venous and small vessel circulation contributes to mortality among COVID-19 patients.(*7*) This coagulopathy is thought to be due to pro-inflammatory factors.(*8*) However, given the high rates of antiphospholipid antibodies among COVID-19 patients, this thrombosis could also be promoted by autoimmunity.(*9*) In other diseases (e.g., Epstein-Barr, parvovirus B19, and hepatitis B or C viruses), it is known that post-infectious autoantibodies can occur, even transiently, and cause a substantial increase in disease severity, including post-viral thrombosis.(*10, 11*)

The goal of this study was to investigate the possibility that COVID-19 patients have autoimmune antibodies to Annexin A2, a critical protective and anti-inflammatory protein expressed in the lung.(*12, 13*) Annexin A2 has an important role in fibrinolysis, cell membrane stabilization and repair, and maintaining the integrity of the pulmonary microvasculature.(*14, 15*) Anti-Annexin A2 antibodies are known to be associated with a higher rate of thrombotic events among patients with antiphospholipid disorders.(*16*) Given the important protective role of Annexin A2 in the lung, we performed ELISAs on plasma obtained from hospitalized COVID-19 patients to determine whether any antibodies were directed against Annexin A2. For comparison, we also studied antibodies directed against Annexin A5, which is another target of prothrombotic antiphospholipid antibodies, but has not been shown to have a direct role in maintaining the integrity of the pulmonary microvasculature.(*17*)

## METHODS

### Study Population

Patient plasma was obtained from 86 hospitalized COVID-19 patients at NYU Langone Health. All of these samples were obtained on hospital day 0 or 1. All patients were confirmed to be COVID-19 positive based on PCR testing. Patients had consented to use of their biospecimens for COVID-19 research through a central biorepository with de-identified clinical data and a protocol approved by the Institutional Review Board at NYU Langone Health.

### ELISA Antibody Testing

Anti-Annexin A2 and anti-Annexin A5 antibodies were measured by ELISA as follows. High binding microwell plates (Immulon 2HB, Thermo Fisher Scientific) were coated overnight at 4°C with 10 μg/mL of human recombinant Annexin A2 or A5 produced in HEK293 cells (Ray Biotech) in phosphate-buffered saline (PBS). After three washes with 0.05% Tween-20 in PBS, the plates were blocked with 0.3% gelatin in PBS at 37°C for 3 hours. The ELISA plates were then washed three times, and plasma samples (1:100 dilution) in 0.1% gelatin in PBS were added to the corresponding wells for 2 hours at 37°C. After washing, bound antibody was detected with anti-human IgG-HRP (Invitrogen). Incubation with the substrate solution for 4 minutes led to color development. The reaction was terminated with the stop solution (Biolegend), and the absorbance was recorded in a microplate reader (VICTOR X2 2030 Multilabel Reader, PerkinElmer, Waltham, Massachusetts) at 450 nm. Relative units (RU) were calculated using a plasma sample previously identified as high responder for IgG autoantibodies. A control ELISA using *E. coli*-derived human Annexin A2 (R&D Systems) was performed as above to confirm that the observed anti-Annexin A2 antibody levels were not due to binding to other human antigens. Testing of 39 patient samples from different severity groups in parallel in *E.coli* or HEK293 cells produced Annexin A2 ELISAs that showed similar results, with an average variation between groups of only 0.17 RU.

### Disease Severity and Laboratory Values

Patients were categorized as 1) non-critical if they were hospitalized, but did not require any critical care treatment, 2) critical ill if they required critical care treatment, or 3) died if they suffered inpatient mortality. Data was also obtained for common laboratory values collected among COVID-19 patients including white blood cell count (WBC), aspartate aminotransferase (AST), alanine aminotransferase (ALT), creatine kinase (CK), lactate dehydrogenase (LDH), Creactive protein (CRP), ferritin, and D-dimer. These values were obtained as the maximum values (i.e., the highest level for each laboratory value over the duration of the patient’s hospitalization).

### Statistical Analysis

We performed descriptive statistics to characterize the study population in terms of age, sex, and race (as categorized by White, Black, Asian or Other). These factors were compared between patients who died and those who survived using rank-sum tests for median age and chi-squared tests for sex, race, and comorbidities. To analyze the maximum laboratory values and the anti-Annexin A2 and A5 antibody levels as stratified by disease severity, we used ANOVA to test for differences in the average values. We also used Kruskal-Wallis tests for differences in the median values of anti-Annexin A2 and A5 antibody levels.

To perform our primary analysis, we tested the association between antibody levels and death using multivariable logistic regression. To account for patient factors, we included age as a continuous variable, sex and race as categorical variables in the regression analysis. We also used diagnosis codes to identify patients with a history of hypertension, diabetes, and obesity (body-mass-index > 30 kg/m^2^) in order to adjust for the most common medical comorbidities among severe COVID-19 patients. A p-value of 0.025 was used to account for the two regression analyses based on a Bonferroni correction. We then used a margins analysis to graphically display mortality risk at a range of antibody levels.

To test the robustness of the association between antibody levels and death, we also performed a sensitivity analysis that added the maximum laboratory values over the course of the hospitalization of these COVID-19 patients. This analysis was performed in order to determine whether the anti-Annexin A2 or A5 antibody levels were an independent predictor of inpatient mortality. As an exploratory analysis, we also evaluated whether antibody levels were associated with any of the maximum laboratory values in an analysis of pairwise correlations. All statistical analyses were performed in Stata 16.2.

## RESULTS

### Study Population Included Hospitalized COVID-19 Positive Patients

Our study population included 86 hospitalized PCR positive COVID-19 patients, of which 28 were non-critical, 36 were critically ill, and 22 had died. Patients that died had higher rates of hypertension (p-value = 0.04) and obesity (p-value = 0.05) when compared to patients who survived. In analyzing the maximum laboratory values among the hospitalized COVID-19 patients, the average values for each lab value increased as expected with disease severity (p-values < 0.01). (See Table 1).

**Table 1:** COVID-19 Study Population Characteristics.

| **Patient Characteristics** | **Hospitalized but Non-Critical (n = 28)** | **Hospitalized and Critically Ill (n = 36)** | **Hospitalized Patients who Died (n = 22)** |
| --- | --- | --- | --- |
| Age |  |  |  |
| Median | 69 | 64 | 69 |
| Range | 28 to 89 | 20 to 101 | 59 to 82 |
| Sex |  |  |  |
| Male | 75% | 75% | 64% |
| Female | 25% | 25% | 36% |
| Race |  |  |  |
| White | 54% | 64% | 54% |
| Black | 18% | 8% | 23% |
| Asian | 7% | 6% | 5% |
| Other | 21% | 22% | 18% |
| Comorbidities |  |  |  |
| Hypertension | 68% | 58% | 86% |
| Diabetes | 32% | 36% | 41% |
| Obesity | 29% | 33% | 55% |
| Maximum Laboratory Values (SD) |  |  |  |
| WBC | 8.9<br>(4.1) | 17.9<br>(11.4) | 20.7<br>(12.0) |
| AST | 62<br>(67) | 193<br>(236) | 461<br>(702) |
| ALT | 63<br>(78) | 160<br>(123) | 268<br>(376) |
| CK | 238<br>(381) | 628<br>(796) | 1249<br>(1909) |
| LDH | 439<br>(318) | 658<br>(333) | 1018<br>(692) |
| CRP | 89<br>(68) | 220<br>(97) | 291<br>(108) |
| Ferritin | 1221<br>(1561) | 3809<br>(6633) | 7439<br>(10947) |
| D-dimer | 736<br>(695) | 4088<br>(3634) | 4954<br>(3658) |

### Levels of Anti-Annexin A2 Antibodies, but not Anti-Annexin A5 Antibodies, Correspond with Disease Severity

Stratified by disease severity, we found higher average levels of anti-Annexin A2 IgG antibodies among hospitalized COVID-19 patients that died when compared non-critical hospitalized COVID-19 patients (p-value = 0.006) and critically ill hospitalized COVID-19 patients (p-value = 0.04). We also found that the median anti-Annexin A2 IgG levels were statistically different when stratified by disease severity (p = 0.01). In comparison, there was no statistically significant difference in the average or median levels of anti-Annexin A5 IgG antibodies when stratified by disease severity (p-values of 0.32 and 0.29). (Figure 1).

**Figure 1:**
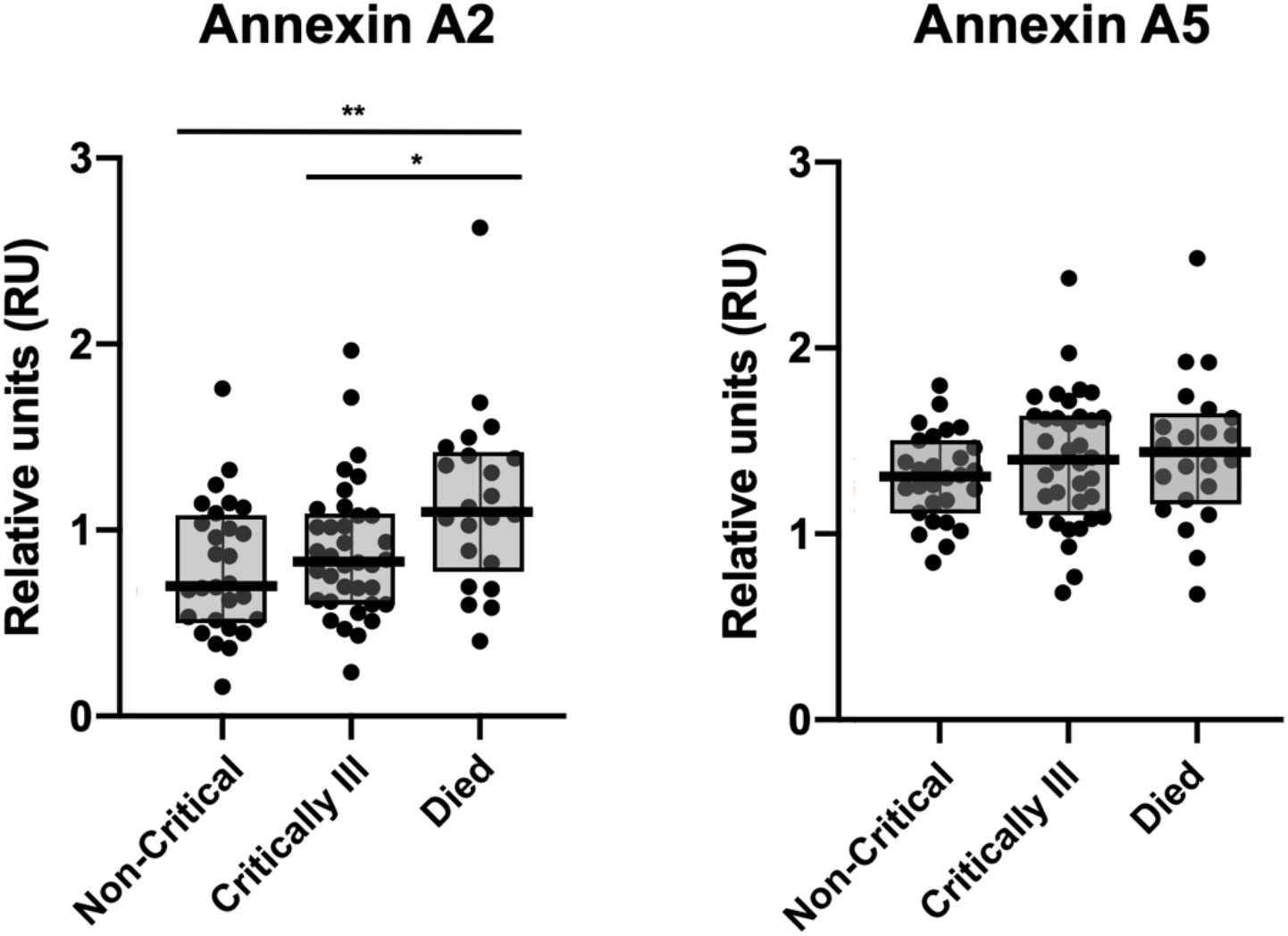
Anti-Annexin A2 and A5 Antibody Levels by COVID-19 Disease Severity. Levels of anti-Annexin A2 and A5 antibodies stratified by disease severity. Measured in relative units (RU). Interquartile ranges (gray shading) and medians (black bars). Statistically significant differences noted as: *p-value < 0.05 and **p-value < 0.01.

### Levels of Anti-Annexin A2 Antibodies, but not Anti-Annexin A5 Antibodies, Predict Mortality After Adjustment for Age, Sex, Race, and Comorbidities

In our primary analysis of mortality among the 86 hospitalized COVID-19 patients in the study population, we found that anti-Annexin A2 antibody levels strongly predicted death after adjustment for age, sex, race, and comorbidities with an odds ratio of 9.3 per RU (95% CI: 1.9 to 44.6, p = 0.005). In comparison, anti-Annexin A5 antibody levels were not associated with a higher mortality rate (95% CI: 0.5 to 15.2, p = 0.22). Using a margins analysis, we graphically depicted predicted mortality rates across a range of levels for anti-Annexin A2 and A5 antibodies and only anti-Annexin A2 antibody levels significantly predicted mortality (Figure 2).

**Figure 2:**
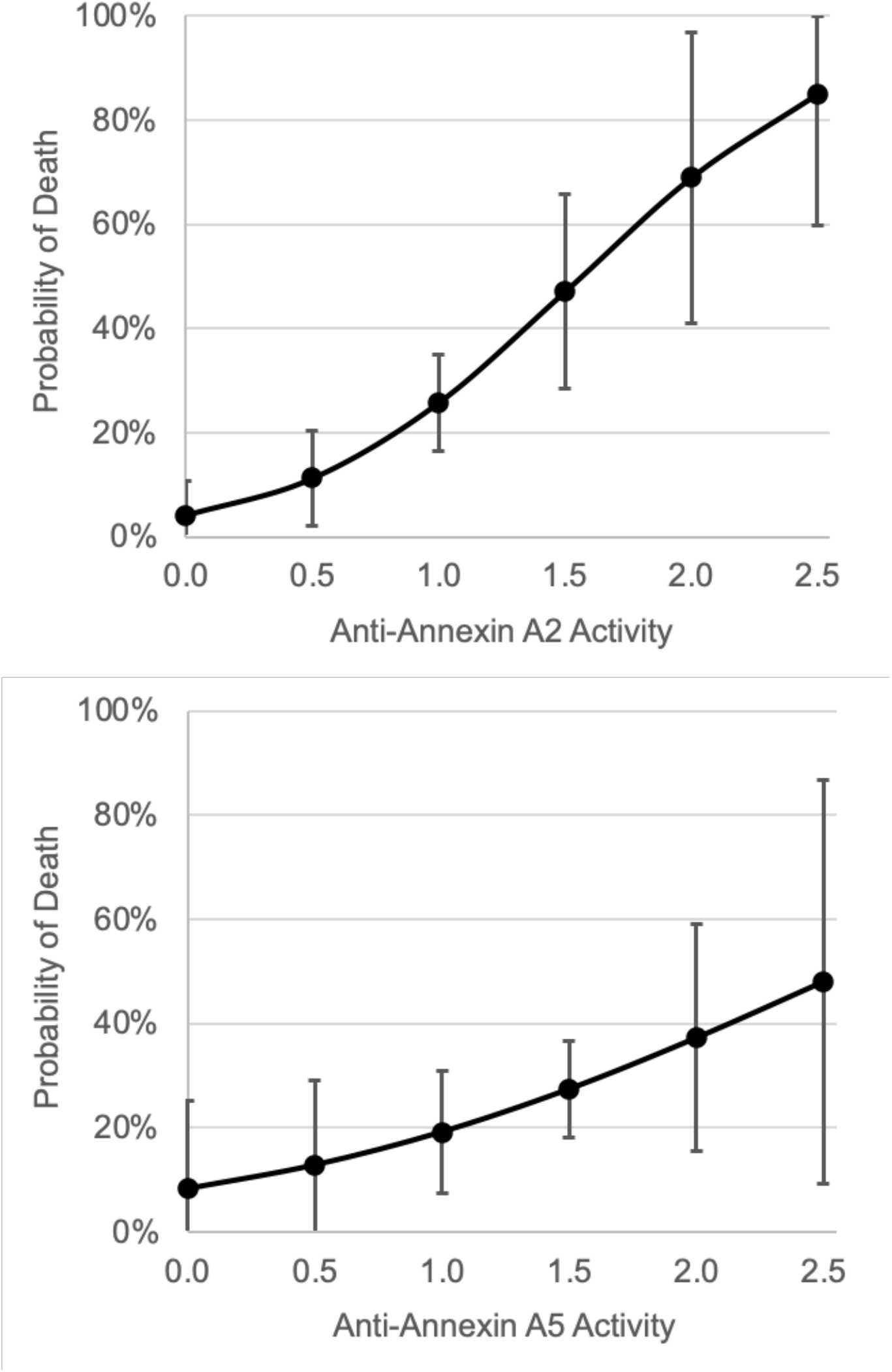
Prediction of Mortality Based on Anti-Annexin A2 and A5 Antibody Levels. Margins analysis based on logistic regression results depicts predicted mortality across a range of anti-Annexin A2 and A5 antibody levels in relative units (RU). Error bars depict 95% confidence intervals.

### Anti-Annexin A2 Antibody Levels are an Independent Predictor of Mortality among Hospitalized COVID-19 Patients

In our sensitivity analysis, we added adjustments for the maximum laboratory values for WBC, AST, ALT, CK, LDH, CRP, ferritin and D-dimer levels to the analyses. We found that anti-Annexin A2 antibody levels still strongly predicted mortality with an odds ratio of 12.9 per RU (95% CI: 1.5 to 108.7, p = 0.019). In comparison, anti-Annexin A5 antibody levels again were not associated with a higher mortality rate (95% CI: 0.4 to 35.5, p = 0.29). (See Supplemental Materials for full regression results).

### Antibody Levels at Initial Hospitalization Correlate with Maximum Abnormalities of Key Laboratory Markers of Severe COVID-19

We also analyzed which of the maximum laboratory values during the hospitalization of COVID-19 patients were associated with the anti-Annexin A2 or A5 IgG antibody levels as measured at hospital day 0 or 1. In this analysis, we found that anti-Annexin A2 antibody levels were correlated with maximum ALT and maximum CRP values over the hospitalization of these COVID-19 patients with correlation coefficients of 0.24 (p = 0.02) and 0.27 (p = 0.01) respectively. As for anti-Annexin A5 antibody levels, we found that they were correlated with maximum WBC with a correlation coefficient of 0.22 (p = 0.04). (Table 2).

**Table 2:** Correlation of Antibody Levels with Other Laboratory Abnormalities.

| Maximum Laboratory Values | Anti-Annexin A2 Antibody Levels |  | Anti-Annexin A5 Antibody Levels |  |
| --- | --- | --- | --- | --- |
|  | Correlation | Significance | Correlation | Significance |
| WBC | 0.14 | 0.20 | **0.22** | **0.04** |
| AST | 0.12 | 0.28 | -0.07 | 0.55 |
| ALT | **0.24** | **0.02** | 0.00 | 0.98 |
| CK | 0.12 | 0.30 | 0.00 | 0.98 |
| LDH | 0.15 | 0.17 | 0.02 | 0.85 |
| CRP | **0.27** | **0.01** | 0.10 | 0.34 |
| Ferritin | 0.17 | 0.12 | -0.01 | 0.94 |
| D-Dimer | 0.09 | 0.42 | 0.18 | 0.10 |

## DISCUSSION

Our study finds evidence of higher levels of IgG antibodies directed against Annexin A2 among COVID-19 patients who died. Although the comparisons in anti-Annexin A2 antibody levels as stratified by disease severity were not vastly different in terms of magnitude, the differences between fatal cases and non-critical or critically ill cases were statistically significant. More importantly, anti-Annexin A2 antibody levels strongly predicted mortality after controlling for patient risk factors and for the maximum levels of key laboratory markers associated with severe COVID-19. We did not identify similar findings for Annexin A5, another target of antiphospholipid antibodies known to be associated with thrombosis.

Previously, the pathophysiology of severe COVID-19 had largely been attributed to a cytokine storm, but this hypothesis has been questioned given that the cytokine levels in COVID-19 are not as high as would be expected in severe lung injury.(*18*) Therefore, the underlying cause of severe COVID-19 still remains to be explained.(*5*) Several recently published studies have found that autoimmunity may play a key role in the pathophysiology of COVID-19, including the presence of autoantibodies against type I interferons in a subset of critically ill patients.(*19, 20*) Another important study demonstrated that a high proportion of hospitalized COVID-19 patients had commonly tested antiphospholipid antibodies detected (i.e., anti-cardiolipin, anti-ß2 glycoprotein, and anti-phosphatidylserine/prothrombin antibodies).(*3*)

In analyzing the response of the adaptive immune system, a pivotal study demonstrated high levels of extrafollicular B cell activation among severe COVID-19 patients.(*21*) Though these patients developed high titers of anti-spike SARS-CoV-2 antibodies, they still experienced poor outcomes.(*21*) Other studies have also shown higher levels of anti-spike antibodies (but not anti-nucleocapsid antibodies) among severe COVID-19 patients when compared to cases that recovered, but these findings have largely been attributed as a response to higher viral loads among severe patients.(*22*) While this explanation is plausible, some studies have suggested that viral load may actually be similar when comparing asymptomatic and symptomatic COVID-19 cases.(*23-25*) In addition, there seems to be a mismatch between the time when the SARS-CoV-2 virus can be cultured from the respiratory tract (in the first week of illness) and the onset of severe respiratory distress (in the second week of illness).(*26*) The timing of these clinical manifestations may be critically important in defining the pathophysiology of severe COVID-19.

Some studies of COVID-19 suggest that certain anti-spike IgG antibodies could be pathogenic, stimulating hyper-inflammatory macrophages, disrupting the pulmonary endothelial barrier, and inducing microvascular thrombosis.(*27*) These findings recapitulate the same findings from studies of the SARS-CoV-1 virus.(*28*) The results of our study may also relate to other studies of SARS-CoV-1, which demonstrated that the humoral responses to the novel coronavirus could result in autoantibodies to important self-antigens in the lung.(*29, 30*) Antibodies generated in response to the second domain of the spike protein of SARS-CoV-1 induced a cytotoxic cross-reactivity to lung epithelial and endothelial cells, specifically targeting Annexin A2.(*29, 30*)

Autopsy evidence demonstrates that COVID-19 patients have extensive thrombotic disease, diffuse alveolar damage, and endothelial disruption that leads to pulmonary edema and fibrin deposition.(*6*) These findings correlate to the clinical manifestations of severe COVID-19, which are diffuse clotting, adult respiratory distress syndrome (ARDS), non-cardiogenic pulmonary edema, and fibrinous pulmonary exudates without bacterial superinfection.(*5*) In the vasculature, Annexin A2 participates in fibrinolysis by complexing with S100A10, forming a co-receptor for tissue plasminogen activator and plasminogen, generating plasmin and promoting fibrin clearance.(*31, 32*) Among patients with antiphospholipid disorders, the presence of Annexin A2 autoantibodies is strongly associated with higher rates of arterial, venous, and small vessel thrombosis, which have been observed in COVID-19.(*33, 34*)

More recently, it was also demonstrated that Annexin A2 supports the integrity of the vascular endothelium by keeping cell junctions tight, especially in response to hypoxia.(*35*) Therefore, inhibition of Annexin A2 might cause non-cardiogenic pulmonary edema and may explain the profound hypoxia, which is a defining characteristic of hospitalized COVID-19 patients. Annexin A2 is also expressed in pulmonary epithelial cells and is involved in cell membrane stabilization and repair.(*12*) Its inhibition can induce apoptosis and loss of lung tissue elasticity.(*36*) Therefore, antagonism of Annexin A2 might explain the severe lung damage that occurs in COVID-19, which can result in ARDS and respiratory failure.

While antagonism of Annexin A2 could explain the hallmark clinical findings of COVID-19 patients, the differences in anti-Annexin A2 antibody levels, though statistically significant, were not very different in terms of magnitude. This finding may be due to the inherent limitations of plasma determinations, which only measure circulating antibody levels and may underestimate the amount of antibody bound to tissues.(*37*) Cross-reactive binding of SARS-CoV-2 antibodies to self-antigens may also be relatively weak and difficult to measure. Furthermore, patient factors such as age and medical comorbidities may determine which patients are able to survive an autoimmune insult.(*38*) In our regression analyses, we found that anti-Annexin A2 antibody levels were strongly associated with mortality after controlling for patient factors. Even when we included the maximum laboratory abnormalities during the hospitalization of these COVID-19 patients, we found that anti-Annexin A2 antibody levels were an independent predictor of death.

Though we do not present direct evidence of the pathogenicity of these anti-Annexin A2 antibodies in severe COVID-19, our study should spur further investigation into the role of autoimmunity induced by the SARS-CoV-2 virus. The presence of these autoantibodies has often thought to be non-pathogenic given that some patients may have high levels of circulating autoantibodies without any evidence of disease.(*39, 40*) Another explanation may be that these autoantibodies are only markers of some other underlying disease process that occurs in COVID-19. However, a failure of immune tolerance may in part explain the substantial variation in outcomes after exposure to SARS-CoV-2.(*38*) Like many other coronaviruses, SARS-CoV-2 may induce a mild to moderate viral syndrome or cause no symptoms at all. However, other patients die from COVID-19 even without any prior medical history or other risk factors.

In this subset of severe patients, the immune system may fail to distinguish certain epitopes of the novel virus from self-antigens, inducing a transient autoimmunity that may cause critical illness.(*38*) The clinical course of severe COVID-19 has been characterized by a second phase of the disease that occurs around 7 to 10 days after onset of symptoms.(*5*) The timing of this respiratory distress appears to coincide with the development of the adaptive immune response.(*41*) In addition, an initial autoimmune injury can cause cellular damage, which in turn, may lead to epitope spreading and autoimmunity to other self-antigens.(*42, 43*) If any of these autoimmune phenomena become more chronic, they may explain the long-term symptoms that some patients have experienced.(*4*)

Our study signals the need to emphasize further research on the role of autoimmunity in COVID-19, especially to protective pulmonary and vascular proteins. We note that several animal models of COVID-19 (as in SARS and MERS) focus on the first phase of disease, which is clearly virally-mediated.(*44, 45*) However, many of these animals do not die from exposure to these emerging novel coronaviruses or develop the severe manifestations found in humans, which in COVID-19, may involve an immune-mediated thrombosis that leads to specific pathologic findings.(*45*) In addition, clinical trials directed at treating severe COVID-19 patients may need to include interventions used in other catastrophic autoimmune diseases especially if autoimmunity is found to have an important role in the pathophysiology of COVID-19.(*46, 47*)

### Limitations

Our study population was obtained from patients at NYU Langone Health, which has three general acute care hospitals across New York City and Long Island. Samples obtained from COVID-19 patients at our institution may not be generalizable to other COVID-19 patients given that patients in this region were seen earlier in the first wave of the pandemic and the characteristics of the SARS-CoV-2 virus may have changed over time. Our study also only included an analysis of hospitalized COVID-19 patients, additional studies should evaluate whether there are differences in the levels of these autoantibodies when compared to patients who were asymptomatic or not hospitalized. Finally, the associations identified in this study cannot be taken to be evidence of causation. Further research is needed to demonstrate whether these anti-Annexin A2 antibodies are pathogenic and have a direct role in the pathophysiology of COVID-19.

## Data Availability

Data will not be made available at this time.

## ACKNOWLEDGMENTS

This research was supported by an internal grant from the NYU Langone COVID-19 Special Fund. We want to specifically thank the Center for Biospecimen Research and Development at the NYU School of Medicine, Brian Fallon for bioinformatics support, and all of the volunteers that helped to obtain and process the samples used in this study and other research related to COVID-19.

## CONTRIBUTIONS

A.R. and D.C.L. designed the study. M.Z. and C.G. performed the experiments. A.R. and P.C. collected the data and identified patient samples at NYU Langone Health. D.C.L. drafted the manuscript and performed the statistical analyses. All authors interpreted the data and provided critical input to the manuscript.

## SUPPLEMENTAL MATERIALS

**Prediction of Mortality Based on Anti-Annexin A2 Antibody Levels After Adjustment for Age, Sex, Race and Comorbidities**

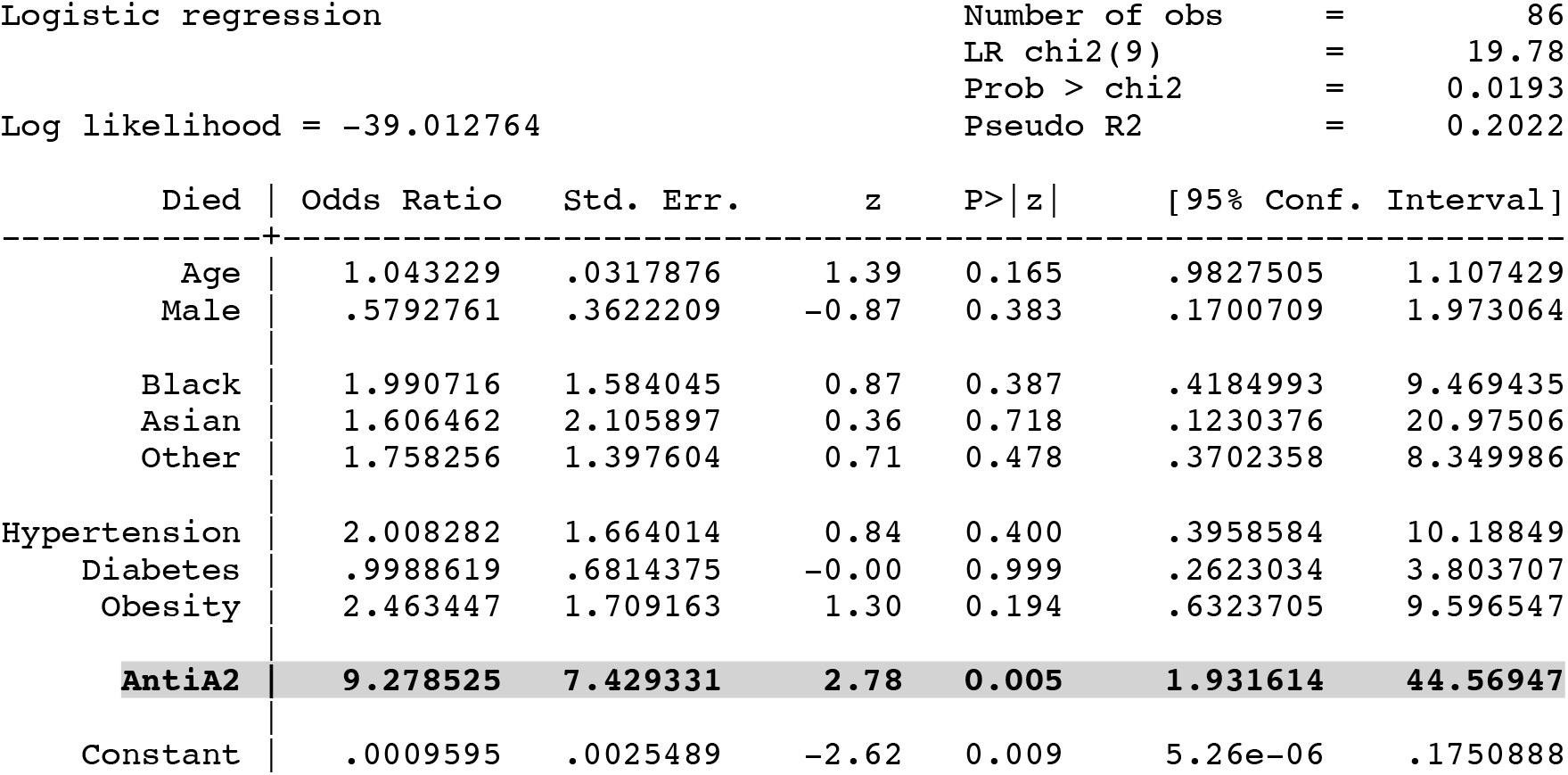

**Prediction of Mortality Based on Anti-Annexin A2 Antibody Levels After Additional Adjustment for Maximum Lab Values**

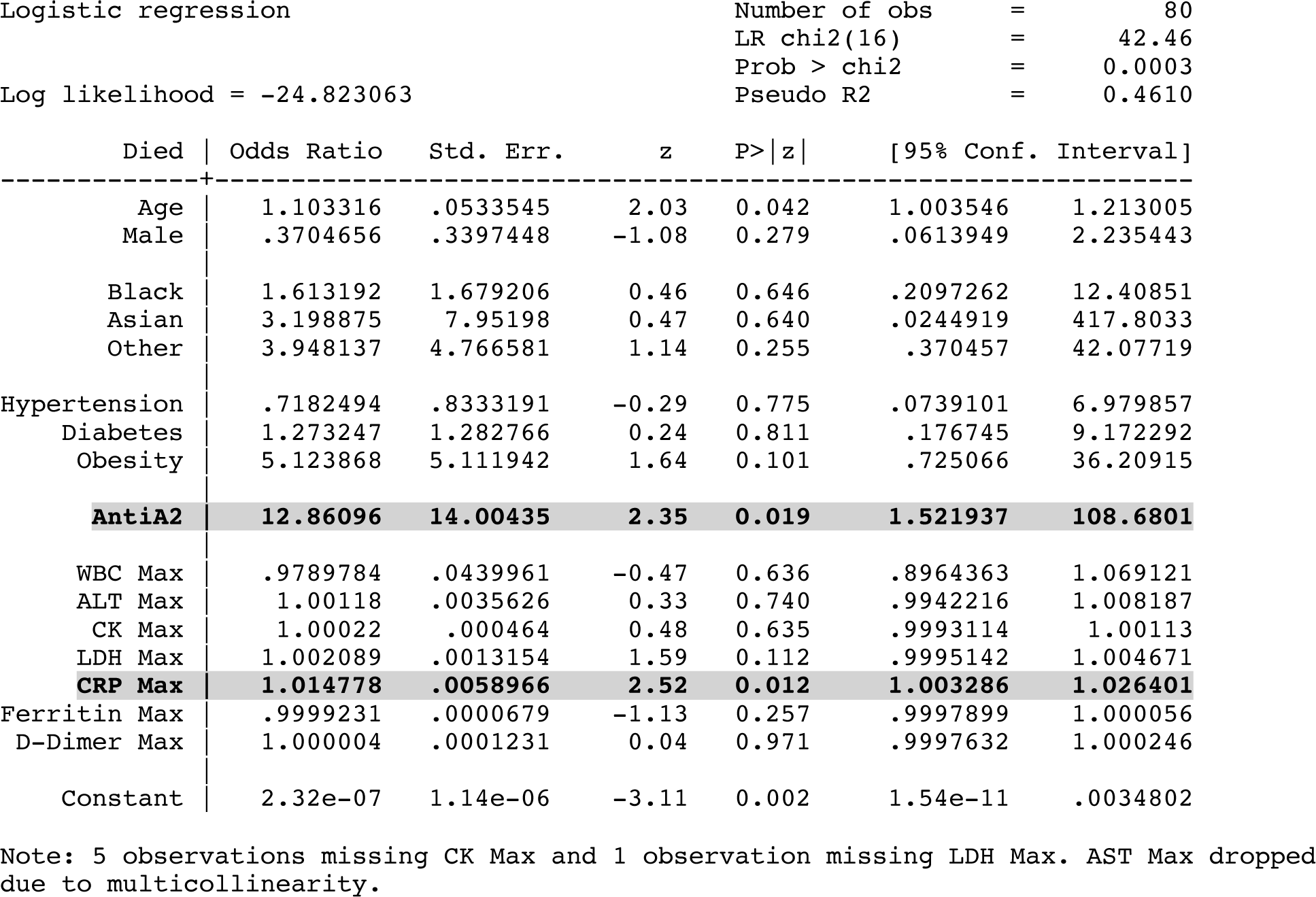

**Prediction of Mortality Based on Anti-Annexin A5 Antibody Levels After Adjustment for Age, Sex, Race and Comorbidities**

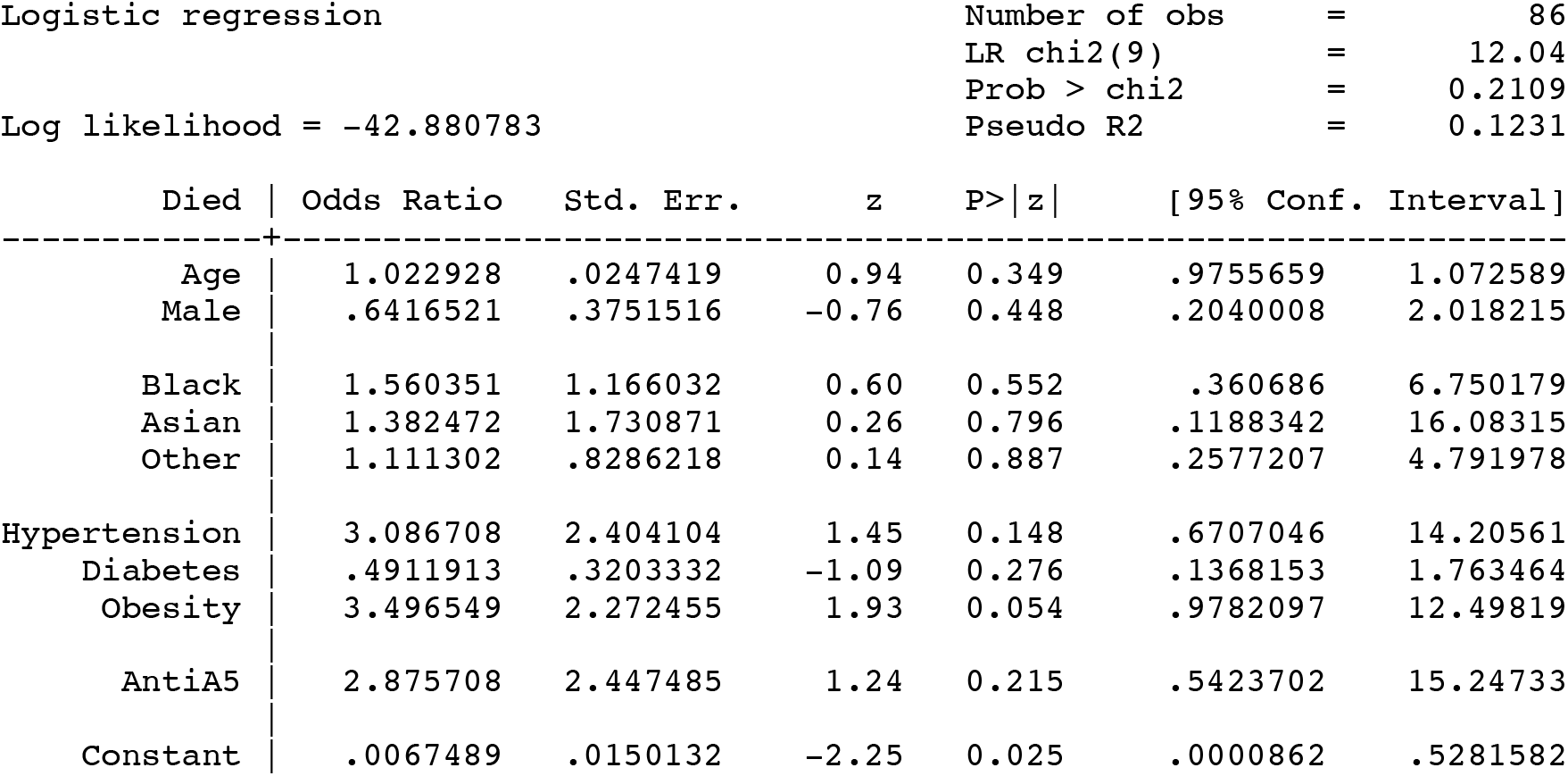

**Prediction of Mortality Based on Anti-Annexin A5 Antibody Levels After Additional Adjustment for Maximum Lab Values**

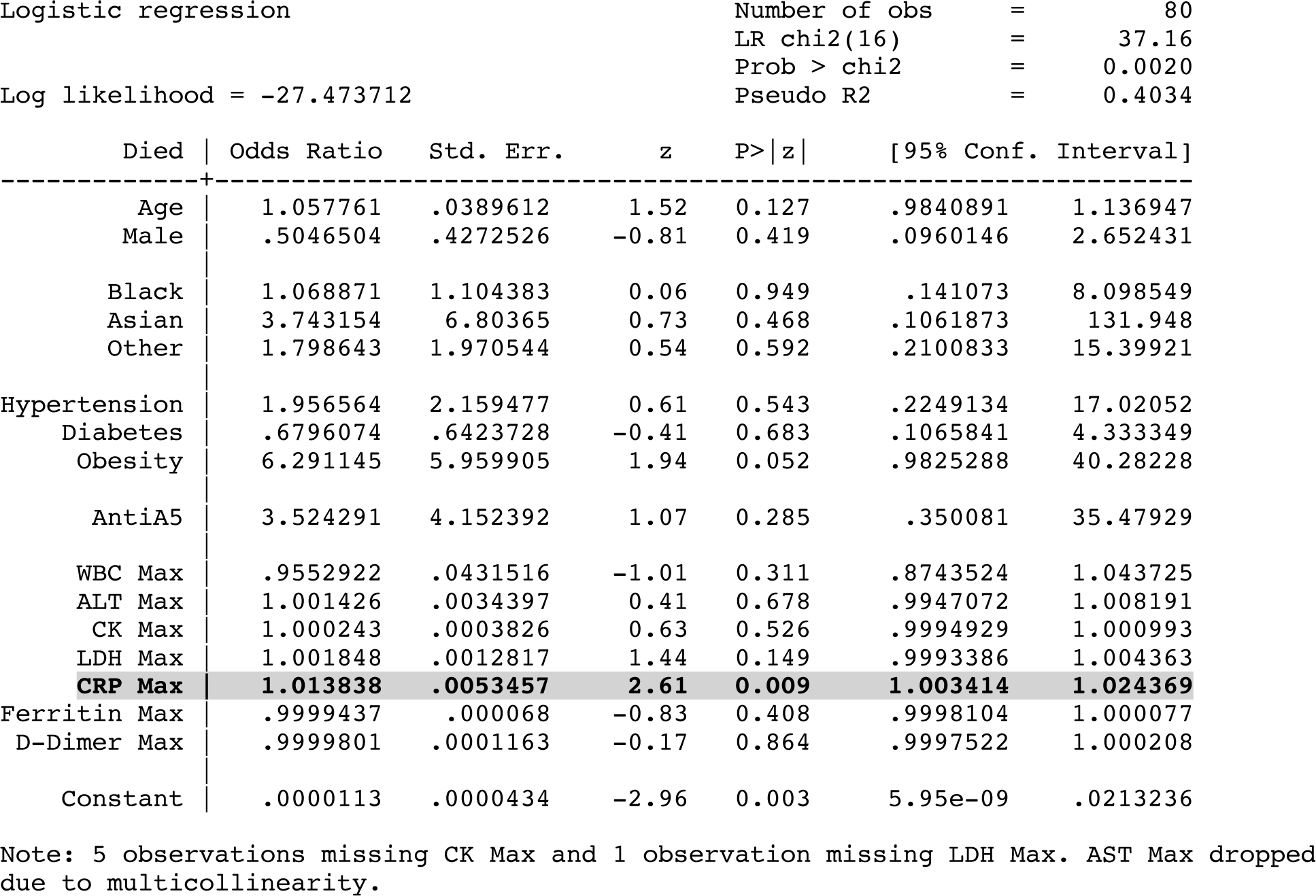

## REFERENCES

1. P. Sinha, M. A. Matthay, C. S. Calfee, Is a “Cytokine Storm” Relevant to COVID-19? JAMA Intern Med 180, 1152–1154 (2020).

2. C. Galeotti, J. Bayry, Autoimmune and inflammatory diseases following COVID-19. Nat Rev Rheumatol 16, 413–414 (2020).

3. Y. Zuo et al., Prothrombotic autoantibodies in serum from patients hospitalized with COVID-19. Sci Transl Med 12, (2020).

4. A. Carfì, R. Bernabei, F. Landi, Persistent Symptoms in Patients After Acute COVID-19. Jama 324, 603–605 (2020).

5. W. J. Wiersinga, A. Rhodes, A. C. Cheng, S. J. Peacock, H. C. Prescott, Pathophysiology, Transmission, Diagnosis, and Treatment of Coronavirus Disease 2019 (COVID-19): A Review. Jama 324, 782–793 (2020).

6. A. V. Rapkiewicz et al., Megakaryocytes and platelet-fibrin thrombi characterize multiorgan thrombosis at autopsy in COVID-19: A case series. EClinicalMedicine 24, 100434 (2020).

7. S. Bilaloglu et al., Thrombosis in Hospitalized Patients With COVID-19 in a New York City Health System. Jama 324, 799–801 (2020).

8. M. Merad, J. C. Martin, Pathological inflammation in patients with COVID-19: a key role for monocytes and macrophages. Nat Rev Immunol 20, 355–362 (2020).

9. Y. Rodríguez et al., Autoinflammatory and autoimmune conditions at the crossroad of COVID-19. J Autoimmun 114, 102506 (2020).

10. J. Rivera-Correa et al., Autoantibody levels are associated with acute kidney injury, anemia and post-discharge morbidity and mortality in Ugandan children with severe malaria. Sci Rep 9, 14940 (2019).

11. N. Abdel-Wahab, S. Talathi, M. A. Lopez-Olivo, M. E. Suarez-Almazor, Risk of developing antiphospholipid antibodies following viral infection: a systematic review and meta-analysis. Lupus 27, 572–583 (2018).

12. M. Dassah et al., Annexin A2 mediates secretion of collagen VI, pulmonary elasticity and apoptosis of bronchial epithelial cells. J Cell Sci 127, 828–844 (2014).

13. V. Dallacasagrande, K. A. Hajjar, Annexin A2 in Inflammation and Host Defense. Cells 9, (2020).

14. K. A. Hajjar, The Biology of Annexin A2: From Vascular Fibrinolysis to Innate Immunity. Trans Am Clin Climatol Assoc 126, 144–155 (2015).

15. M. Luo, K. A. Hajjar, Annexin A2 system in human biology: cell surface and beyond. Semin Thromb Hemost 39, 338–346 (2013).

16. W. Ao, H. Zheng, X. W. Chen, Y. Shen, C. D. Yang, Anti-annexin II antibody is associated with thrombosis and/or pregnancy morbidity in antiphospholipid syndrome and systemic lupus erythematosus with thrombosis. Rheumatol Int 31, 865–869 (2011).

17. J. H. Rand, X. X. Wu, A. S. Quinn, D. J. Taatjes, The annexin A5-mediated pathogenic mechanism in the antiphospholipid syndrome: role in pregnancy losses and thrombosis. Lupus 19, 460–469 (2010).

18. M. Kox, N. J. B. Waalders, E. J. Kooistra, J. Gerretsen, P. Pickkers, Cytokine Levels in Critically Ill Patients With COVID-19 and Other Conditions. Jama 324, 1565–1567 (2020).

19. P. Bastard et al., Autoantibodies against type I IFNs in patients with life-threatening COVID-19. Science 370, (2020).

20. E. Y. Wang et al., Diverse Functional Autoantibodies in Patients with COVID-19. medRxiv, 2020.2012.2010.20247205 (2020).

21. M. C. Woodruff et al., Extrafollicular B cell responses correlate with neutralizing antibodies and morbidity in COVID-19. Nat Immunol 21, 1506–1516 (2020).

22. S. P. Weisberg et al., Distinct antibody responses to SARS-CoV-2 in children and adults across the COVID-19 clinical spectrum. Nat Immunol, (2020).

23. S. Lee et al., Clinical Course and Molecular Viral Shedding Among Asymptomatic and Symptomatic Patients With SARS-CoV-2 Infection in a Community Treatment Center in the Republic of Korea. JAMA Intern Med 180, 1–6 (2020).

24. S. H. Ra et al., Upper respiratory viral load in asymptomatic individuals and mildly symptomatic patients with SARS-CoV-2 infection. Thorax, (2020).

25. I. Hasanoglu et al., Higher viral loads in asymptomatic COVID-19 patients might be the invisible part of the iceberg. Infection, 1-10 (2020).

26. M. Cevik et al., SARS-CoV-2, SARS-CoV, and MERS-CoV viral load dynamics, duration of viral shedding, and infectiousness: a systematic review and meta-analysis. The Lancet Microbe.

27. W. Hoepel et al., Anti-SARS-CoV-2 IgG from severely ill COVID-19 patients promotes macrophage hyper-inflammatory responses. bioRxiv, 2020.2007.2013.190140 (2020).

28. L. Liu et al., Anti-spike IgG causes severe acute lung injury by skewing macrophage responses during acute SARS-CoV infection. JCI Insight 4, (2019).

29. Y. S. Lin et al., Antibody to severe acute respiratory syndrome (SARS)-associated coronavirus spike protein domain 2 cross-reacts with lung epithelial cells and causes cytotoxicity. Clin Exp Immunol 141, 500–508 (2005).

30. Y. T. Fang et al., Annexin A2 on lung epithelial cell surface is recognized by severe acute respiratory syndrome-associated coronavirus spike domain 2 antibodies. Mol Immunol 47, 1000–1009 (2010).

31. B. Huang et al., Hypoxia-inducible factor-1 drives annexin A2 system-mediated perivascular fibrin clearance in oxygen-induced retinopathy in mice. Blood 118, 2918–2929 (2011).

32. M. Valapala, S. I. Thamake, J. K. Vishwanatha, A competitive hexapeptide inhibitor of annexin A2 prevents hypoxia-induced angiogenic events. J Cell Sci 124, 1453–1464 (2011).

33. F. Cañas, L. Simonin, F. Couturaud, Y. Renaudineau, Annexin A2 autoantibodies in thrombosis and autoimmune diseases. Thromb Res 135, 226–230 (2015).

34. E. Cockrell, R. G. Espinola, K. R. McCrae, Annexin A2: biology and relevance to the antiphospholipid syndrome. Lupus 17, 943–951 (2008).

35. M. Luo et al., Annexin A2 supports pulmonary microvascular integrity by linking vascular endothelial cadherin and protein tyrosine phosphatases. J Exp Med 214, 2535–2545 (2017).

36. S. L. Jiang, D. Y. Pan, C. Gu, H. F. Qin, S. H. Zhao, Annexin A2 silencing enhances apoptosis of human umbilical vein endothelial cells in vitro. Asian Pac J Trop Med 8, 952–957 (2015).

37. F. Tedesco et al., Pathogenic Role of Complement in Antiphospholipid Syndrome and Therapeutic Implications. Front Immunol 9, 1388 (2018).

38. A. N. Theofilopoulos, D. H. Kono, R. Baccala, The multiple pathways to autoimmunity. Nat Immunol 18, 716–724 (2017).

39. J. Land, A. Rutgers, C. G. Kallenberg, Anti-neutrophil cytoplasmic autoantibody pathogenicity revisited: pathogenic versus non-pathogenic anti-neutrophil cytoplasmic autoantibody. Nephrol Dial Transplant 29, 739–745 (2014).

40. R. I. Litvinov et al., Distinct specificity and single-molecule kinetics characterize the interaction of pathogenic and non-pathogenic antibodies against platelet factor 4-heparin complexes with platelet factor 4. J Biol Chem 288, 33060–33070 (2013).

41. N. Sethuraman, S. S. Jeremiah, A. Ryo, Interpreting Diagnostic Tests for SARS-CoV-2. Jama 323, 2249–2251 (2020).

42. D. Salem et al., β2-Glycoprotein I-specific T cells are associated with epitope spread to lupus-related autoantibodies. J Biol Chem 290, 5543–5555 (2015).

43. U. S. Deshmukh, H. Bagavant, J. Lewis, F. Gaskin, S. M. Fu, Epitope spreading within lupus-associated ribonucleoprotein antigens. Clin Immunol 117, 112–120 (2005).

44. T. C. Sutton, K. Subbarao, Development of animal models against emerging coronaviruses: From SARS to MERS coronavirus. Virology 479-480, 247-258 (2015).

45. S. N. Ehaideb, M. L. Abdullah, B. Abuyassin, A. Bouchama, Evidence of a wide gap between COVID-19 in humans and animal models: a systematic review. Crit Care 24, 594 (2020).

46. A. D. Truong et al., Therapeutic plasma exchange for COVID-19-associated hyperviscosity. Transfusion, (2020).

47. P. Tabarsi et al., Evaluating the effects of Intravenous Immunoglobulin (IVIg) on the management of severe COVID-19 cases: A randomized controlled trial. Int Immunopharmacol, 107205 (2020).

